# Efficacy and safety of Desidustat versus Erythropoietin in post kidney transplant anaemia: a prospective observational study

**DOI:** 10.1101/2025.03.03.25322925

**Authors:** Smita Divyaveer, Pooja Barak, Sahil Kharbanda, Prasanna Ethiraj S, Vignesh Subramani, Rajesh Kumar, Kushal Kekan, Madhuri Kashyap, Kanchan Prajapati, Deepy Zohmangaihi, Nabhajit Mallik, Deepesh Lad, Madhumita Premkumar, Rahul Yadav, Raja Ramachandran, Harbir Singh Kohli

## Abstract

**Background:** Post-renal transplant anemia significantly impacts patient outcomes and quality of life. The only option available until recently was erythropoietin (EPO) that requires intravenous or subcutaneous injection. New class of drugs that are Hypoxia-Inducible Factor Prolyl Hydroxylase Inhibitor (HIF PHI) have become available and there is scarce data on their use in kidney transplant recipients (KTRs).

**Methods:** This was prospective observational study which included 58 renal transplant recipients with anemia (Hb < 10 g/dL) who were started on either oral HIF PHI in the form of Desidustat (n=30) or EPO (n=28) based on nephrologist discretion. Adult KTRs with a hemoglobin (Hb) level of less than 10 gm/dl were included. Patients with acute graft dysfunction, bleeding and primary haematological diseases were excluded. Baseline clinical characteristics were recorded. Primary outcomes were change in Hb at 8 weeks in the two groups. Secondary outcomes included inflammatory markers (CRP, ESR), iron indices, and safety.

**Results:** 58 patients were enrolled in the study; 30 received desidustat and 28 received erythropoietin. Both groups showed significant improvement in Hb from baseline to 2 months (Desidustat: 8.69 ± 0.86 g/dL to 9.89 ± 0.92 g/dL; EPO: 8.54 ± 0.73 g/dL to 9.55 ± 0.71 g/dL; p > 0.05). No significant differences were observed in inflammatory markers or iron indices. Both treatments were well-tolerated, with no reported adverse events.

**Conclusions:** Desidustat demonstrated comparable efficacy and safety to EPO, with the added benefit of oral administration. It presents a promising alternative for anemia management in post-renal transplant patients.

## Introduction

Anemia is a frequently encountered complication among patients after renal transplantation, affecting a substantial proportion during the post-transplant period. Reports on the prevalence of anemia in these patients vary widely, ranging from 20% to 51%, depending on the study. This variation is influenced by factors such as the definitions of anemia and the time elapsed since transplantation^1,2^. The prevalence of anemia in post-transplant patients varies according to time since kidney transplantation. In the early post-transplant period, it can often be attributed to factors like surgical blood loss, reduced erythropoietin production due to allograft dysfunction, and the effects of immunosuppressive medications. Typically, anemia resolves within three to six months in patients with well-functioning allografts^3^. However, six to twelve months post-transplant, other contributors such as iron deficiency, chronic inflammation, graft dysfunction, drugs, the influence of inflammatory markers, infections such as play a more prominent role in the persistence of anemia**^Error!^ ^Reference^ ^source^ ^not^ ^found.^**. Uncorrected anemia not only reduces patients’ quality of life but is also associated with cardiovascular complications^4^, graft failure, and mortality^5^.

Effectively managing anemia in post-renal transplant patients requires understanding its duration and causes. Despite optimisation of immunosuppressive therapy, treatment of infections and nutritional deficiencies, patients with chronic allograft dysfunction continue to have anemia requiring erythropoietin similar to patients with anemia of chronic disease in chronic kidney disease (CKD). While a new class of drugs namely hypoxia-inducible factor prolyl hydroxylase inhibitor (HIF-PHI) have been used to treat the anemia in CKD, data on use of this class of drugs in kidney transplant recipients (KTRs) is scarce. These drugs work by activating the hypoxia-inducible factor pathway, thereby promoting the production of endogenous erythropoietin. Studies conducted in other patient groups, such as those with chronic kidney disease (CKD), have shown that HIF-PHI can lower inflammatory markers like IL-6 and CRP and improve iron metabolism^6^.

Not all HIF PHIs are available equally across the globe. In India, desidustat, a hypoxia-inducible factor prolyl hydroxylase inhibitor (HIF-PHI) is an available alternative treatment option for renal anemia. Given its demonstrated efficacy in raising hemoglobin levels and reducing inflammation in other populations, desidustat offers a dual benefit—correcting anemia while simultaneously addressing the underlying inflammatory burden that often complicates treatment. It is being used as an alternative to erythropoietin in treatment of anemia in KTRs. Therefore, we designed this observational study to evaluate the efficacy and safety of desidustat in the treatment of anemia in kidney transplant recipients (KTRs) and to assess its impact on inflammatory markers in comparison to erythropoietin therapy.

## Materials and Methods

### Study design and population

This single- centre, prospective, observational pilot study was conducted at the outpatient department of the Post Graduate Institute of Medical Education and Research (PGIMER), Chandigarh, Punjab. The study (CTRI/2024/08/072458) was conducted between September 2023 and October 2024. Renal transplant recipients with anemia (Hb < 10 g/dl) were screened. Patients with stable graft function defined as <25% increase in serum creatinine and no change in immunosuppressive medication dosages in prior 3 months were included. Patients with active bleeding, ongoing acute graft on chronic dysfunction, primary hematological disorders, presence of malignancies, pregnant or lactating individuals and on dialysis were excluded. Patients with nutritional deficiencies were supplemented and patients were reassessed after repletion of deficiencies for eligibility. Patients who developed severe infections requiring hospital admission or bleeding from any site during study period were excluded from analysis as these factors can affect hemoglobin levels independently.

### Treatment of anemia

Eligible patients from the outpatient department of Nephrology at PGIMER, Chandigarh were enrolled. Patients started on either erythropoietin or desidustat, as determined by their primary care nephrologist, were assigned to erythropoietin and desidustat groups respectively. The protocol of dosing of erythropoietin and desidustat is given in supplementary figure 1 and 2. Clinical and laboratory data were recorded. Routine investigations for CKD patients with anemia were documented, including complete blood count (CBC), peripheral smear, reticulocyte count, stool occult blood, serum iron studies, B12 levels, and folate levels. Patients were enrolled into erythropoietin or desidustat groups and followed up for 2 months.

### Study assessments

The patients were evaluated for medical history and laboratory investigations, including hemoglobin levels (Hb), serum iron, serum ferritin, total iron-binding capacity (TIBC), and inflammatory markers including erythrocyte sedimentation rate (ESR), C-reactive protein (CRP), levels of tacrolimus or cyclosporine.

Primary outcome measured were changes in Hb levels at 8 weeks. Secondary outcomes focused on changes in inflammatory markers (CRP, ESR), iron indices, cyclosporine or tacrolimus level. Adverse events were recorded as minor if treated on outdoor basis and major if they required admission to hospital.

### Sample size and statistical analysis

This was a pilot observational study, and a feasibility sample size of 60 was decided. This study included 58 renal transplant recipients diagnosed with anaemia, of whom 30 received desidustat and 28 received erythropoietin. The data was collected, and appropriate statistical analysis was performed using epi-info version 7.2. Demographic and baseline characteristics are summarized using descriptive statistics. Categorical variables are summarized with frequency and percentage. Continuous variables are summarized with mean, standard deviation, etc. A P value of Continuous variables were analysed using ANOVA, and categorical data were compared using chi-square tests. A p-value < 0.05 was considered significant. Missing data were assessed with Little’s MCAR test. Multiple imputation was used for MAR data, and sensitivity analyses confirmed consistent results. Analyses were conducted using Epi Info 7.2, with minimal impact on findings due to low missingness (<10%).

## ETHICS STATEMENT

This pilot study was reviewed and approved by the institutional ethics committee (approval number: IEC-INT/2023-SPL1024 A). The study was conducted in accordance with the ethical principles that have their origin in the Declaration of Helsinki and in accordance with the International Conference on Harmonization’s Good Clinical Practice guidelines (ICH-GCP). All participants provided written and informed consent.

## Results

Total of 83 renal transplant recipients with anaemia were screened. 20 patients were excluded for various reasons. All 63 patients completed 2 months of follow up. However, during the study period, 3 patients developed severe infection, 1 had massive gastrointestinal bleeding and 1 patient was non-compliant to medication for more than 2 weeks. These 5 patients were excluded from analysis. Data of 58 patients; 30 in desidustat group and 28 in erythropoietin group were included in analysis. The details of patient recruitment flow chart are shown in figure 1 below. The baseline hemoglobin, iron indices and iron profile were similar in both the study groups. The baseline characteristics of study participants are shown in table 1 below. The mean duration since time of transplantation was 4.01 ± 2.23 years and 3.53 ± 1.98 years. The causes of chronic allograft dysfunction were chronic rejection (n=21), chronic calcineurin toxicity (n= 4), disease recurrence (n=4) and BK Virus nephropathy (n=1) in the desidustat group. The etiologies of chronic graft dysfunction in the erythropoietin group were chronic rejection (n=19), chronic calcineurin toxicity (n= 3), disease recurrence (n=3), non-specific chronicity (n=2) and non-recovery of AKI (n=1). 25 and 22 patients were on triple immunosuppression in desidustat and erythropoietin groups respectively. 11 patients among the entire study population were dual immunosuppression for at least 3 months prior to enrolment (patients with change in immunosuppression within prior 3 months were excluded from the study). In these patients calcineurin inhibitors were stopped other patients in view of calcineurin toxicity or for stepping down immunosuppression for repeated infections. All patients were on anti-proliferative medication; 23 and 22 on Mycophenolate and 7 and 6 patients in each group on Azathioprine in desidustat and erythropoietin groups respectively.

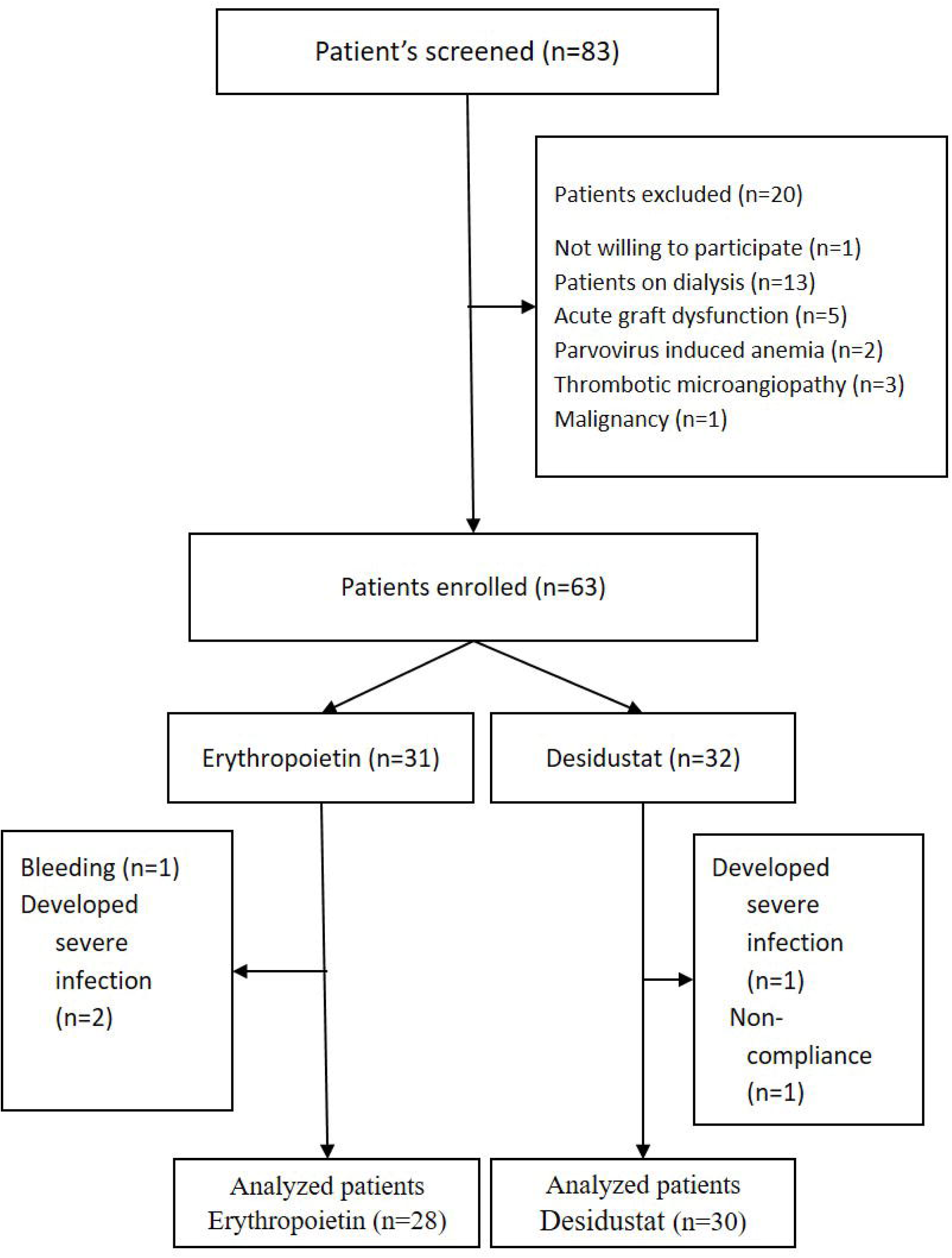

**Table 1:** Baseline characteristics of study participants in the Desidustat and Erythropoietin groups.

|  |  |  |  |
| --- | --- | --- | --- |
| Parameter | Desidustat<br>(n=30)<br>(Mean ± SD) | Erythropoietin<br>(n=28)<br>(Mean ± SD) | p-value |
| Age (years) | 49.7 ± 16.77 | 50.96 ± 16.68 | 0.775 |
| Sex (Male/Female) | 16/14 | 19/9 | 0.195 |
| Creatinine (mg/dL) | 3.44 ± 1.06 | 3.09 ± 0.81 | 0.169 |
| Hemoglobin (g/dL) | 8.69 ± 0.86 | 8.54 ± 0.73 | 0.488 |
| Iron (µg/dL) | 96.55 ± 27.13 | 96.46 ± 27.57 | 0.990 |
| TIBC (µg/dL) | 283.39 ± 58.46 | 306.74 ± 59.54 | 0.137 |
| Transferrin Saturation (%) | 33.38 ± 8.90 | 35.43 ± 8.65 | 0.377 |
| MCV (fL) | 88.30 ± 5.70 | 89.82 ± 6.57 | 0.349 |
| MCH (pg) | 29.13 ± 1.74 | 29.39 ± 1.77 | 0.575 |
| MCHC (g/dL) | 33.50 ± 1.17 | 33.29 ± 1.12 | 0.479 |
| Vitamin B12 (pg/mL) | 400.54 ± 111.51 | 394.25 ± 110.97 | 0.830 |
| Folate (ng/mL) | 12.8 ± 4.83 | 12.45 ± 4.3 | 0.775 |
| LDH (U/L) | 185.45 ± 33.2 | 195.53 ± 32.74 | 0.250 |
| Ferritin (ng/mL) | 316.8 ± 115.45 | 329.75 ± 121.18 | 0.679 |
| CRP (mg/L) | 4.31 ± 1.76 | 4.37 ± 1.94 | 0.911 |
| ESR (mm/hr) | 23.59 ± 8.6 | 27.4 ± 8.34 | 0.094 |
| AST (U/L) | 25.17 ± 8.55 | 24.68 ± 9.4 | 0.837 |
| ALT (U/L) | 26.47 ± 8.96 | 25.68 ± 9.86 | 0.751 |
| Calcium (mg/dL) | 9.52 ± 0.67 | 9.67 ± 0.53 | 0.350 |
| Phosphate (mg/dL) | 3.7 ± 0.61 | 3.48 ± 0.52 | 0.150 |
| Vitamin D (ng/mL) | 64.66 ± 20.21 | 65.02 ± 20.45 | 0.947 |
| iPTH (pg/mL) | 141.83 ± 17.78 | 138.35 ± 15.79 | 0.435 |
| **Abbreviations:** **AST:** Aspartate aminotransferase **ALT:** Alanine aminotransferase<br>**iPTH:** Intact parathyroid hormone **LDH:** Lactate dehydrogenase **CRP:** C-reactive protein<br>**ESR:** Erythrocyte sedimentation rate **TIBC:** Total iron-binding capacity |  |  |  |

### Change in hemoglobin and iron studies in the two study groups

Both groups demonstrated significant within-group increases in hemoglobin levels over 8 weeks. Desidustat group had improvement in Hb from 8.69 ± 0.86 g/dL to 9.89 ± 0.92 g/dL, and erythropoietin group had improvement in Hb from 8.54 ± 0.73 g/dL to 9.55 ± 0.71 g/dL. No significant inter-group differences were observed (p=0.12).[ Figure 2]. At baseline, the mean iron levels were 96.55 ± 27.13 µg/dL for desidustat group and 96.46 ± 27.57 µg/dL for Erythropoietin group (p = 0.990). By week 8, the levels had slightly increased, but no significant inter-group differences were found (p = 0.499). Similar trends were observed for total iron binding capacity (TIBC) and transferrin saturation (Tsat), with no significant changes or group differences over 2 months (all p-values > 0.05).

**Figure 2:**
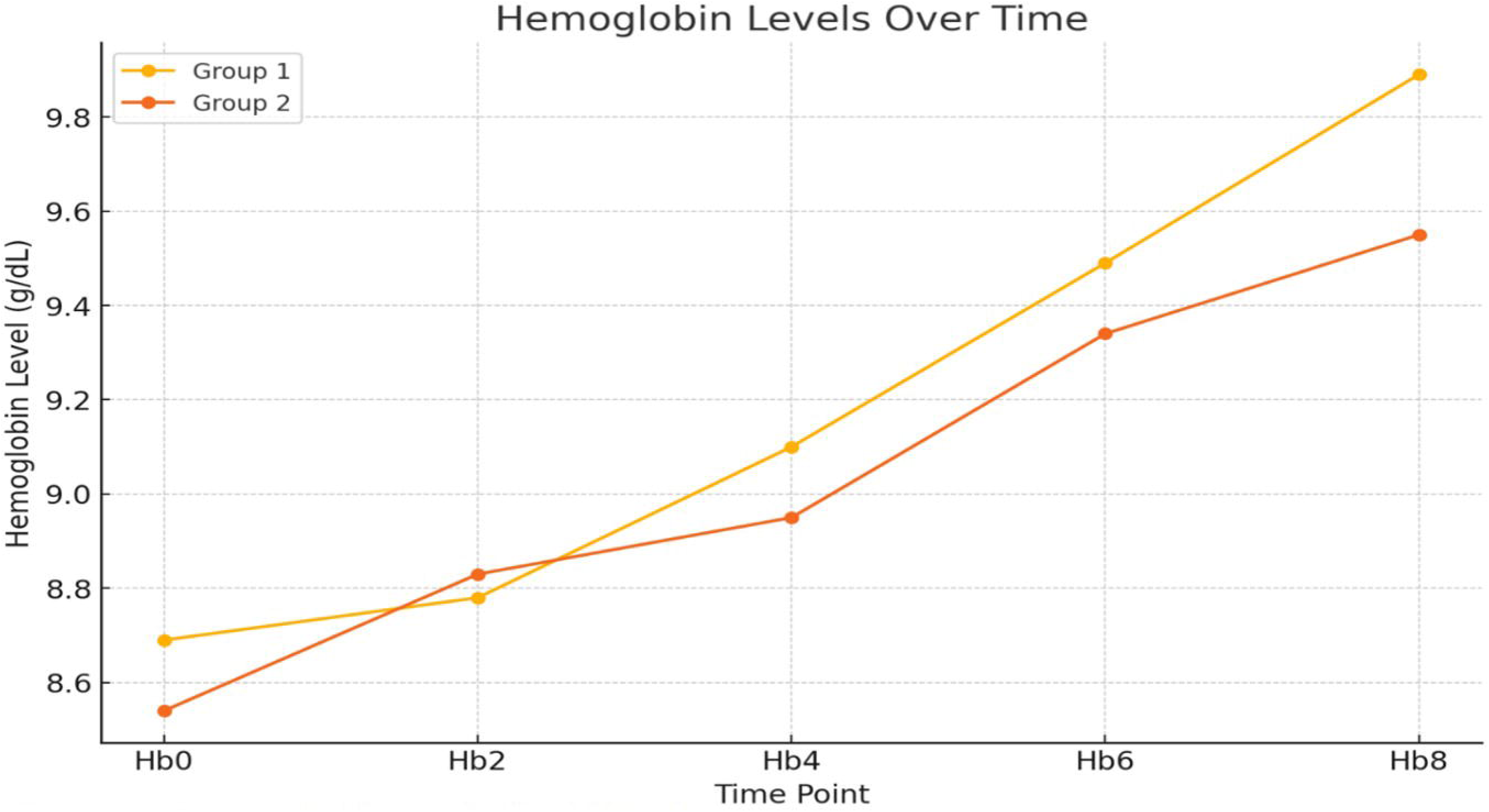
Change in Hemoglobin(Hb) in the two groups. Group 1= Desidustat. Group 2= Erythropoietin; Hb0,Hb2,Hb4,Hb6&Hb8 indicate Hb at 0,2,4,6&8 weeks

### Inflammatory Markers

No significant changes were observed in CRP and ferritin between the groups over the study period (Table 2). ESR was significantly higher in erythropoietin group on follow up as compared to Desidustat group.

**Table 2:** Comparison of inflammatory makers in the Desidustat and erythropoietin groups at baseline and follow up.

| Inflammatory Markers | Baseline |  |  | Follow up (8 weeks) |  |  |
| --- | --- | --- | --- | --- | --- | --- |
|  | Desidustat group | Erythropoietin group | P value | Desidustat group | Erythropoietin group | P value |
| CRP (mg/L) | 4.31 ± 1.76 | 4.37 ± 1.94 | 0.911 | 4.12 ± 1.94 | 4.4 ± 1.35 | 0.528 |
| ESR (mm/hour) | 23.59 ± 8.59 | 27.40 ± 8.34 | 0.094 | 24.36 ± 4.5 | 29.56 ± 7.69 | 0.002 |
| Ferritin (ng/mL) | 316.8 ± 115.45 | 329.75 ± 121.18 | 0.679 | 462.3.0 ± 119.8 | 497.57 ± 112.43 | 0.253 |

Abbreviations: CRP: C reactive protein, ESR: Erythrocyte sedimentation rate

### Cyclosporine and Tacrolimus Levels

In desidustat and erythropoietin groups 23 and 18 patients were on Tacrolimus respectively and 2 and 4 patients were on Cyclosporine respectively. At baseline, the mean tacrolimus levels were 6.10 ± 0.65 ng/mL in desidustat group 1 and 6.02 ± 0.58 ng/mL in erythropoietin group (p = 0.657). At week 8, these levels increased slightly (statistically insignificant) to 6.31 ± 0.69 ng/mL and 6.35 ± 0.53 ng/mL, respectively (p = 0.831). Baseline cyclosporine levels were 83.5 ± 7.78 ng/mL in desidustat group and 79.5 ± 12.76 ng/mL in erythropoietin group (p = 0.714). At week 8, the levels were 96.5 ± 4.95 ng/mL and 99.5 ± 15.29 ng/mL, respectively, with no significant differences observed (p = 0.810).

### Adverse events

3 patients developed severe infection in the form of urinary tract infection, dengue fever and pneumonia during the study period and 1 patient developed massive bleeding requiring blood transfusion due to diverticulosis. The patient who developed urinary tract infection has prior history of multiple urinary tract infections due to bladder abnormality. No other adverse events, either minor or major, were reported in either treatment group during the study period. No patient reported any gastrointestinal intolerance to desidustat. One patient was non-compliant to all medication, not desidustat alone (excluded from analysis).

## Discussion

HIF PHIs are emerging as an alternative to treat anemia in CKD but there is scarce data on use of these agents in kidney transplant recipients (KTRs). Desidustat is approved for use in India and is readily available for use in CKD patients in India owing to its comparable cost and efficacy to erythropoietin. It is used off label in renal transplant recipients and there is no data to the best of our knowledge on use of Desidustat in KTRs. Although this is an observational study, it has largest sample size on use of Desidustat in transplant population to date. This study provides evidence for desidustat’s comparable efficacy to EPO, establishing it as a viable and patient-friendly treatment option.

Desidustat is a novel therapeutic agent belonging to the HIF-PHI class, which replicates the body’s hypoxic response^7^. Under normal oxygen conditions, the HIF-α subunit undergoes hydroxylation and is subsequently degraded. However, HIF-PHIs inhibit this hydroxylation process, stabilizing HIF-α. This allows HIF-α to bind with HIF-β and activate genes responsible for erythropoiesis, iron homeostasis, and angiogenesis^8^. This mechanism not only increases endogenous erythropoietin production but also reduces hepcidin levels, enhancing iron mobilization^9^. These properties make HIF-PHIs particularly effective in treating inflammation-related anemia, which is commonly observed in post-transplant patients. In contrast to ESAs, which act directly on erythroid progenitors to stimulate red blood cell production, HIF-PHIs provide the added advantage of addressing disrupted iron metabolism through hepcidin suppression^10^. Only one prior study prospective randomised controlled study in KTRs has analysed the efficacy of HIF PHI in improving anemia in KTRs after 6 months of kidney transplant^11^. This study reported the efficacy of Roxadustat, however, the control group received iron alone (none of the patients during the study period received erythropoietin) and the degree of anemia was mild at baseline (10.56 ± 0.44 gm/dl). Our study was designed to compare Desidustat with erythropoietin as the latter is the standard of care in patients with CKD and anemia. The degree of baseline anemia was moderate (average 8.6 gm/dl) in our study. We found equivalence of erythropoietin and desidustat in improving Hb in KTRs with a comparable safety profile.

The effect of Desidustat in our study is similar to a prior study by Provenzano et al. in CKD patients that demonstrated HIF-PHIs’ effectiveness in stimulating erythropoiesis^12^ with maintained stable iron indices (serum iron, TIBC, and TSAT) throughout the study, with no significant differences observed. This corroborates with another study by Locatelli et al., which noted that short-term HIF-PHI therapy does not immediately alter iron parameters^13^. However, long-term data, such as that from Li et al., suggests that HIF-PHIs improve iron bioavailability through sustained hepcidin reduction^14^. Studies with longer follow up are warranted to ascertain the effect on Desidustat on Iron profile parameters.

The dual mechanism of improvement in iron metabolism and inflammation exerted by HIF PHIs may offer significant benefits, particularly for individuals who do not respond adequately to traditional ESA therapy due to inflammatory conditions as reported with use of Vadadustat in CKD^15^. Coyne et al. reported similar findings, underscoring the anti-inflammatory effects and iron-modulating capabilities of HIF-PHIs, which are particularly relevant in the post-transplant setting^16^. Some HIF PHIs such as roxadustat^17^ and molidustat^18^ have been reported to have improved anemia especially in states with increased inflammation characterised by raised CRP^19^. But a similar effect was not found with Daprodustat^20^. In our study, the inflammatory markers (CRP and ESR) showed no significant inter-group differences, further supporting the safety desidustat with regards to these inflammatory makers although a superiority of Desidustat in presence of increased inflammation could not be assessed due to a small sample size.

HIF PHIs have had several safety concerns because of which they are not globally used as an alternative to erythropoietin. The major concerns have been with regards to cardiovascular safety, carcinogenic concerns, risk of increased thrombosis^21^ and risk of infections^22^. The adverse effects do not seem to be uniform across all HIF PHIs and hence cannot be considered a class effect. Some agents have been approved in various countries for use in CKD patients eg. Daprodustat and Vadadustat have been approved for use in US. Desidustat was developed in India and has been approved for use in CKD patients with anemia since 2022^23^. Available literature on Desidustat indicates that it is devoid of major adverse effects unlike other drugs of this class. It is being used off label in transplant due to its availability and ease of administration. We found that both desidustat is well-tolerated, with no significant interactions with immunosuppressive agents, such as tacrolimus and cyclosporine. There we 2 severe infections episodes but it cannot be ascertained if Desidustat was contributory. This aligns with Miki et al. findings, which demonstrated that roxadustat, another HIF-PHI, maintains a strong safety profile in post-transplant populations^24^.

Our study lays down the first report of efficacy and safety of Desidustat in KTRs. However, there are some limitations. Although we did not find any rejection episodes during the study, we cannot ascertain the effect of Desidustat on pharmacokinetics of anti-proliferative agents as therapeutic level monitoring is not routinely done for these agents for all patients. We did not perform a comprehensive analysis with regard to anti-inflammatory effect of Desidustat on markers such as interleukin 6 and Hepcidin in our study population. The follow up duration in our study is relatively short and the sample size is small, hence, larger studies with longer follow up are warranted to study the effect of Desidustat on long term outcomes.

## Conclusion

Desidustat is as efficacious as erythropoietin in treatment of anemia in post-renal transplant patients. It has a similar effect on iron profile and inflammatory parameters as compared to erythropoietin. Adverse events were similar between the two groups. Due to its ease of administration, Desidustat can be a patient-centered option in clinical practice for treatment of anemia post kidney transplant. Future research should explore its long-term effects, cost-effectiveness, and broader therapeutic potential.

## Data Availability

All data produced in the present work are contained in the manuscript

## Acknowledgments

The authors thank the study participants without whom the study would not have been completed. Thanks to all the medical and paramedical staff involved in clinical care of all study participants.

