## Supplementary figures and images for "Efficacy and safety of Desidustat versus Erythropoietin in post kidney transplant anaemia: a prospective observational study"

### supplementary figure 1 and 2

**Supplementary material**

Supplementary Figure1

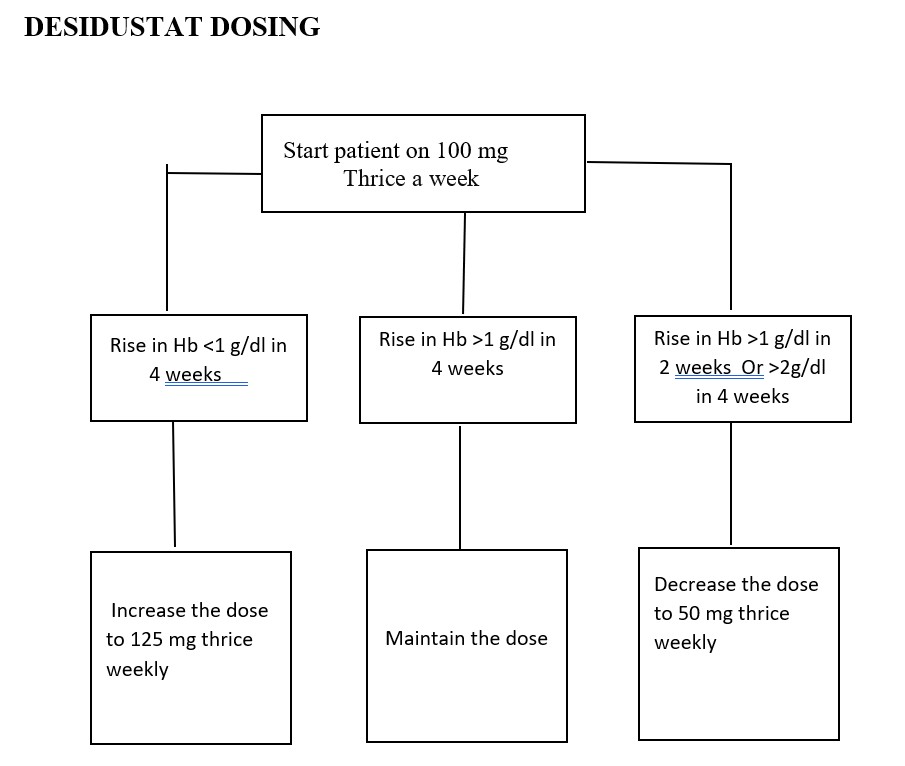

#### Supplementary Figure 2

#### ESA-DOSING

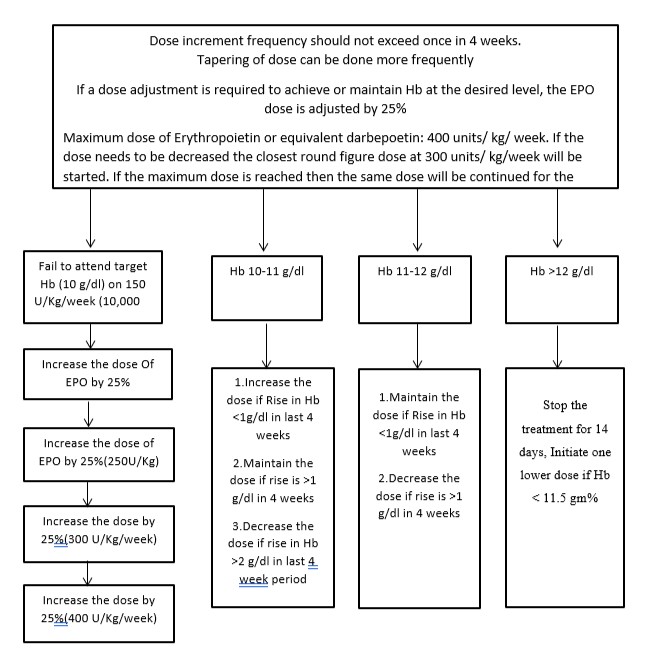
